# Deep resequencing of the 1q22 locus in non-lobar intracerebral hemorrhage

**DOI:** 10.1101/2023.04.18.23288754

**Authors:** Livia Parodi, Mary E Comeau, Marios K Georgakis, Ernst Mayerhofer, Jaeyoon Chung, Guido J Falcone, Rainer Malik, Stacie L Demel, Bradford B Worrall, Sebastian Koch, Fernando D Testai, Steven J Kittner, Jacob L McCauley, Christiana E Hall, Douglas J Mayson, Mitchell SV Elkind, Michael L James, Daniel Woo, Jonathan Rosand, Carl D Langefeld, Christopher D Anderson

## Abstract

**Objective:** Genome-wide association studies have identified *1q22* as a susceptibility locus for cerebral small vessel diseases (CSVDs), including non-lobar intracerebral hemorrhage (ICH) and lacunar stroke. In the present study we performed targeted high-depth sequencing of *1q22* in ICH cases and controls to further characterize this locus and prioritize potential causal mechanisms, which remain unknown.

**Methods:** 95,000 base pairs spanning *1q22*, including *SEMA4A, SLC25A44* and *PMF1*/*PMF1-BGLAP* were sequenced in 1,055 spontaneous ICH cases (534 lobar and 521 non-lobar) and 1,078 controls. Firth regression and RIFT analysis were used to analyze common and rare variants, respectively. Chromatin interaction analyses were performed using Hi-C, ChIP-Seq and ChIA-PET databases. Multivariable Mendelian randomization (MVMR) assessed whether alterations in gene-specific expression relative to regionally co-expressed genes at *1q22* could be causally related to ICH risk.

**Results:** Common and rare variant analyses prioritized variants in *SEMA4A* 5’-UTR and *PMF1* intronic regions, overlapping with active promoter and enhancer regions based on ENCODE annotation. Hi-C data analysis determined that *1q22* is spatially organized in a single chromatin loop and that the genes therein belong to the same Topologically Associating Domain. ChIP-Seq and ChIA-PET data analysis highlighted the presence of long-range interactions between the *SEMA4A*-promoter and *PMF1*-enhancer regions prioritized by association testing. MVMR analyses demonstrated that *PMF1* overexpression could be causally related to non-lobar ICH risk.

**Interpretation:** Altered promoter-enhancer interactions leading to *PMF1* overexpression, potentially dysregulating polyamine catabolism, could explain demonstrated associations with non-lobar ICH risk at *1q22*, offering a potential new target for prevention of ICH and CSVD.

## Introduction

Stroke is the second leading cause of death and first cause of adult disability worldwide.^1^ Despite substantial advances in therapy and disease prevention, the risk of lifetime stroke continues to rise.^1^ Accounting for approximately 10-40% of strokes,^2^ spontaneous (non-traumatic) intracerebral hemorrhages (ICH) are responsible for 50% of stroke-related mortality.^3^ Caused by rupture of small penetrating vessels, ICH can be classified as lobar or non-lobar, depending on the location of the bleeding.^3^ Lobar ICH, affecting the cerebral cortex or cortical-subcortical junction, is predominantly associated with cerebral amyloid angiopathy (CAA). Non-lobar ICH originates in deep structures of the cerebral hemisphere, brainstem, and cerebellum, and is associated with hypertension and other vascular risk factors, coexisting white matter disease, and silent infarctions.^3^

Genetic risk factors are estimated to account for 30% of ICH risk.^4^ The exploration of this genetic background could be crucial to the identification of pathways potentially targetable by novel therapeutic strategies. Large multicenter collaborations developed to advance the investigation of the genetic drivers of ICH culminated in the first large multicenter Genome-Wide Association Study (GWAS) that detected a group of variants at the *1q22* locus in association with increased risk of non-lobar ICH.^5^ The *1q22* locus spans a 95 kb region of strong linkage disequilibrium harboring four genes, *SEMA4A, SLC25A44, PMF1* and *PMF1-BGLAP*, the latter coding for the natural read-through product between *PMF1* and the neighboring gene *BGLAP*.

Subsequent GWAS analyses of traits pathophysiologically related to ICH rediscovered variants at *1q22* in association with small vessel ischemic strokes,^6^ and with white matter hyperintensities,^7,8^ extending the contribution of *1q22* to susceptibility to other manifestations of cerebral small vessel disease (CSVD).

While GWAS analyses are powerful tools to identify regions associated with a trait of interest, they often highlight groups of variants in areas of linkage disequilibrium (LD), prohibiting prioritization of specific causal variants at single-variant resolution.^9^ Building upon previous GWAS findings,^5–8^ in the present study we sought to further characterize the *1q22* locus with the goal of prioritizing potential causal mechanisms for further biological exploration. After assembling a cohort of 2,133 ICH patients and healthy controls, we completed deep sequencing of the 95 kb region spanning the locus and analyzed the resulting data combining a variety of synergistic methods aimed at delving into the genetic, epigenetic, and transcriptional background of this susceptibility locus.

## Methods

**Figure 1** summarizes the analytical workflow adopted in the present study. We assembled a cohort of 1,055 spontaneous ICH cases (534 lobar and 521 non-lobar) and 1,078 controls (**Fig 1A**), which were deep sequenced across the *1q22* locus by the Northwest Genomics Center at the University of Washington (**Fig 1B**). Firth regression, RIFT analysis and fine mapping analyses were performed to detect and prioritize both common and rare single variants contributing to ICH risk at this region (**Fig 1C**). Publicly available Hi-C, ChIP-Seq and ChIA-PET data were used to further investigate chromatin organization and to detect the presence of long-range interactions within the *1q22* locus (**Fig 1D**). Finally, multivariable Mendelian randomization analysis was computed to assess the causal relationship between alteration in expression of any of the *1q22* genes and risk of ICH (**Fig 1E**).

**Figure 1.**
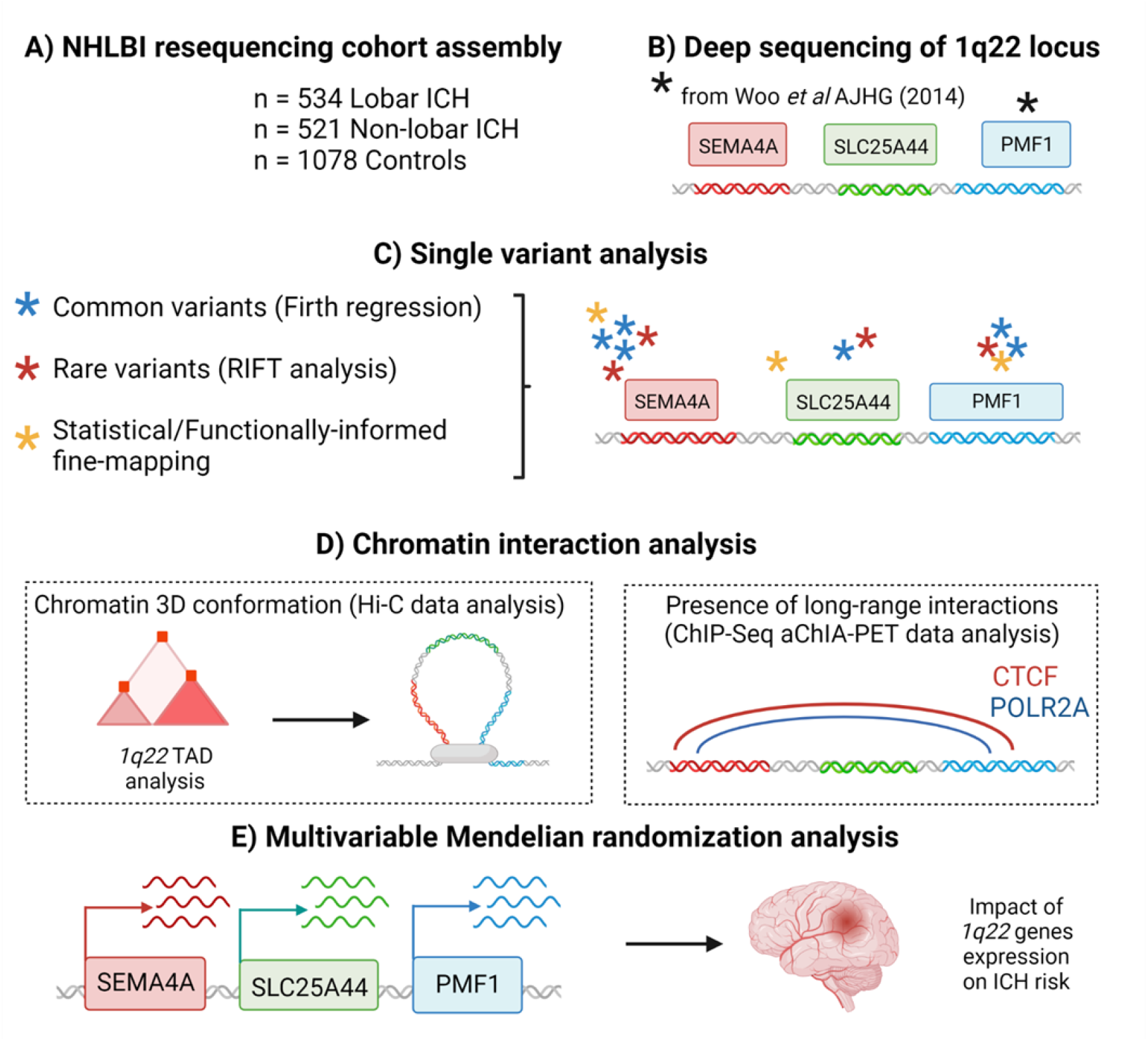
Overview of the analytical workflow adopted in the present study. A cohort of 1,055 ICH patients and 1,078 controls was recruited (A) and submitted to deep sequencing targeting locus *1q22*, a susceptibility locus for non-lobar ICH previously discovered by Woo *et al*.^5^ A black asterisk indicates the genomic location of the GWAS top hits (B). Resulting data were analyzed leveraging multiple approaches to assess the impact of single variants (C), and to understand *1q22* 3D chromatin conformation (D). Finally, multivariable Mendelian randomization analysis was performed to understand whether dysregulation of *1q22* gene expression could be causally related to the higher ICH risk associated with this locus (E).

### 1. Study cohort

We included 1,055 cases and 1,078 controls recruited through the “Genes and Outcomes of Cerebral Hemorrhage on Anticoagulation” (GOCHA) and “Ethnic/Racial variation in Intracerebral Hemorrhage” (ERICH) studies.^10,11^ Enrollment criteria for both cases and controls were harmonized across the participating institutions; we note that age enrollment requirements did vary, with age > 55 years for GOCHA and > 18 years for ERICH. Only patients of European ancestry (self-reported and genetically verified by genotype data) were included in the present study. ICH location and case status were verified based on centralized adjudication of CT scans at patient presentation. Based on these criteria, 534 lobar ICH and 521 non-lobar ICH cases were identified for inclusion. Controls (n = 1,078) were enrolled from the same geographic region as the cases using random digit dialing (ERICH) and ambulatory clinics (GOCHA), and matched cases by age (+/- 5 years), sex, ethnicity, and race. 18 ICH patients and 176 controls overlap with those included in prior ICH GWAS analyses.^5^ An overview of the cohort is presented in **Table 1**.

**Table 1.** Cohort overview. Age at event/recruitment, relative standard deviation (sd) and age range, and sex proportions are reported for all the ICH patients and controls included in the present study.

|  | Age, mean (sd;<br>range) | Sex (F) |
| --- | --- | --- |
| Lobar (n=534) | 73 (12.6; 21-100) | 51% (272/534) |
| Non lobar<br>(n=521) | 69 (13.3; 29-98) | 39% (205/521) |
| Controls<br>(n=1078) | 70 (12.3; 21-95) | 45% (484/1078) |

### 2. Study approval and patients consent

As stated in GOCHA and ERICH study protocols,^10,11^ all recruiting sites received IRB approval for enrollment. Informed consent was obtained from each participant or legally authorized representative.

### 3. Targeted resequencing of the *1q22* locus

Resequencing services were provided by the Northwest Genomics Center at the University of Washington, Department of Genome Sciences. The *1q22* region was sequenced in all samples, focusing on a 95,000 bp region (chr1: 156,118,869-156,213,964), encompassing *SEMA4A, SLC25A44*, and *PMF1*/*PMF1-BGLAP* genes, using the 96plex Nimblegen SeqCap platform, without fingerprint. Following sequencing, BAM files generated through the Picard data-processing pipeline (http://broadinstitute.github.io/picard) were aligned to GRCh37 human genome reference. IndelRealigner and Base Recalibration tools, provided by Genome Analysis Toolkit (GATK),^12^ were used prior to variant calling to remove duplicates and to locate and realign indels. Finally, GATK HaplotypeCaller (v3.2) was used to jointly call samples, detecting single-nucleotide variants (SNVs), as well as insertions or deletions.

### 4. Quality control

GATK Variant Quality Score Recalibration (VQSR) method was used to retain variants. Only SNVs/indels having a depth >= 10 were included. Variants with call rate < 0.98, case-control call-rate difference > 0.005 were excluded from the present analysis. Samples presenting with low average call rate (< 0.98), low mean sequence depth (< 30), low mean genotype quality (< 85), differential missingness between cases and controls (P < 0.05) and P < 10^−6^ at Hardy-Weinberg Equilibrium (HWE) test, were excluded from the analysis. To avoid any possible bias due to lack of coverage of the sequenced region, we used Samtools (http://www.htslib.org) --depth option on BAM files.

### 5. Single variant analyses

#### 5.1. Firth regression

Single variant tests of association were performed using Firth regression (--glm firth), available through PLINK2.0 software (https://www.cog-genomics.org/plink/2.0/). Resulting data were analyzed first using a standard case-control approach (lobar vs controls and non-lobar vs controls), after retaining only frequent variants (MAF > 1%). As a secondary analysis, because *1q22* is a susceptibility locus for non-lobar ICH alone, non-lobar ICH patients (n = 521) were compared with the pool of both lobar ICH patients and ICH-free controls together (n = 1,612). All regressions were adjusted for age and sex. Additional sensitivity analyses were performed including the first four principal components generated using smartpca and EIGENSTRAT method to account for the presence of population structure. Resulting p-values and the absolute value of the natural logarithm of the odd ratios were used to compare our results with those obtained in previous GWAS analysis.^5^ Wald Z-scores were used in subsequent fine-mapping analyses.

#### 5.2. Rare variant analysis

As an additional method to prioritize variants at *1q22*, we used the Rare Variant Influential Filtering Tool (RIFT),^13^ a newly developed method that uses a leave-one-out strategy to evaluate the impact of each rare variant on the burden test (SKAT-O) results. Results are summarized as the change in the chi square statistics from the aggregate test. Variants were considered as outliers (key influential rare variants in the aggregate test) only if the results of outliers’ analysis converged for all the three methods used (Fence, Tukey, median absolute deviation).

#### 5.3. Fine-mapping analysis

We leveraged multiple fine-mapping tools in our exploration of the *1q22* locus. PAINTOR^14^ was used to perform functionally informed fine-mapping. In the present analysis, variants were annotated using the Python utility “AnnotateLocus.py” and its annotation library that leverages functional data such as those generated by FANTOM5 consortium^15^ and the RoadMap project.^16^ Statistical fine mapping was performed using FINEMAP,^17^ CAVIAR^18^ and SuSiE R package (https://github.com/stephenslab/susieR). These tools differ in terms of how posterior inclusion probabilities (PIPs) are estimated. FINEMAP uses a Shotgun Stochastic Search (SSS) algorithm that, after exploring a large number of possible causal configurations, assigns the highest PIPs to those with non-negligible probability. CAVIAR identifies the minimal set of variants that has the highest probability of containing the causal variant(s), after accounting for the conditional distribution of all association statistics in the locus. SuSiE takes advantage of an iterative Bayesian stepwise selection (IBSS) model to produce a series of minimal credible sets harboring highly correlated variants.

Fine-mapping analyses were computed using the Wald statistic scores generated as previously described and an LD matrix computed using the Python script “CalcLD_1kg_vcf.py”, part of PAINTOR framework, taking advantage of the 1000 Genomes (Phase 3) latest release as reference^19^ and selecting only individuals of European ancestry. The resulting LD matrix, composed of pairwise Pearson correlation coefficients for each SNP, was used as input for all the fine-mapping tools in the present analysis.

#### 5.4. Functional annotation

Publicly available expression data generated in blood by the eQTLGen Consortium^20^ were downloaded to assess whether any of the variants prioritized by single variant analyses could act as eQTLs (i.e., variants correlated with varying levels of gene expression), potentially altering *1q22* genes expression.

ChIP-Seq assay data released as part of the ENCODE project^21^ were used to further characterize gene expression regulation at *1q22*. Chromatin immunoprecipitation followed by sequencing (ChIP-Seq) allows detection of interactions between DNA and specific proteins of interest.^22^ We focused on the presence and interactions involving active promoters and enhancers, as well as on the detection of regions enriched in transcription factor binding sites. We accessed signal data relative to H3K27ac and H3K4me2 histone marks measured in four ICH-relevant cell types (GM12878, H1-hESC and HUVEC) from the UCSC Genome Browser portal (https://genome.ucsc.edu/). Histone modifications such as H3K27ac and H3K4me2 are indicative of how accessible chromatin is at a specific region. In particular, H3K27ac marks are detected in proximity of active promoter and enhancer regions,^23^ while H3K4me2 signal is enriched near transcription factor binding sites.^24^ As a final step, Ensembl database (https://grch37.ensembl.org/index.html) was queried to retrieve the precise location of promoters and enhancers within *1q22*.

### 6. Chromatin interaction analyses

We further pursued our dissection of the *1q22* locus by analyzing its chromatin conformation leveraging ChIP-Seq, Hi-C, and ChIA-PET data. Hi-C, the high throughput version of chromosome conformation capture technique, facilitates mapping of the chromatin folding patterns across the genome, capturing its 3D hierarchical organization and subdividing the genome into Topologically Associated Domains (TADs) by identifying regions physically interacting to form chromatin loops.^25^ We used Juicebox,^26^ a tool that uses publicly available Hi-C data derived from GM12878 cells ^25^ to interactively explore the genome 3D conformation and identify TADs. Along with TAD identification, we used Juicebox to detect chromatin loops, normally visualized in Hi-C maps as intense “peak” pixels that represent areas in which contact frequency is enhanced compared with neighboring regions.^25^ Chromatin peaks are usually located at the corners of contact domains, in proximity to convergent CTCF-binding motifs.^26^ The CCCTC-binding factor, CTCF, is one of the major mediators of DNA loop formation and plays a central role in both chromatin spatial organization and consequent gene expression regulation.^27^

To explore DNA-protein interactions, we combined ChIP-Seq and ChIA-PET data analysis. The ChIA-PET method improves the resolution of DNA-protein and DNA-DNA interactions over ChIP-Seq alone.^28^ ChIP-Seq and ChIA-PET data targeted CTCF and RNA-polymerase II A (POLR2A), another essential component of genes transcription processes.^29^ CTCF-mediated long-range interactions and consequent DNA loop formation, followed by POLR2A recruitment, is crucial for transcription initiation.^30^ Both ChIP-Seq and ChIA-PET data measured in K562 cells, a multipotential leukemia cell line of human origin^31^ and released by the ENCODE Consortium, were downloaded from UCSC Genome Browser portal to complete these analyses.

### 7. Multivariable Mendelian randomization analyses

Multivariable Mendelian randomization (MVMR) analyses were performed to explore whether variations of the expression levels of any of the genes within the *1q22* locus could be causal for higher risk of lobar or non-lobar ICH. After accounting for the possibility that one variant could be simultaneously associated with more than one exposure, MVMR allows estimation of the causal effects of each exposure in a single analysis model.^32^ Genetic instruments acting as *cis*-eQTLs and potentially influencing the expression of genes (log2 transformed) within the *1q22* locus in blood were thus selected using the statistically significant (FDR < 0.05) *cis*-eQTL results available through the eQTLGen Consortium data portal.^20^ Only independent variants were retained, defined by “clumping” for LD at *r*^*2*^ < 0.1 using 1000 Genomes Europeans as reference population.^19^ Genetic associations of the selected instruments with non-lobar ICH were derived from the previously published ICH GWAS.^5^ MVMR analyses were performed combining different MR methods^33^ (inverse weighted variance, Lasso, Egger and weighted median) available through the R-package MendelianRandomization (version 0.7.0).

### 8. Data availability

Sequencing data used in this study are available on dbGAP (https://www.ncbi.nlm.nih.gov/gap/; Accession ID: phs000416.v2.p1) and on the CDKP portal (https://cd.hugeamp.org/downloads.html). *Cis*-eQTL data were available on the eQTLGen Consortium portal (https://www.eqtlgen.org). Additional data supporting these findings are available by the authors, upon reasonable request.

## Results

### 1. Coverage analysis highlights regional variation in sequencing depth

Absence of coverage (average read depth close to 0) was detected in an interval between bp 156,139,280 and 156,140,732, located in an intronic region of the *SEMA4A* gene (**Supplementary Figure S1**). Additional information regarding the quality of the alignment in the poorly covered *SEMA4A* region were retrieved from the correspondent Concise Idiosyncratic Gapped Alignment Report (CIGAR) string. Likely related to a polyT repeat, different segments of this small region did not align with the reference sequence in any of the sequenced samples. Low coverage across samples was also detected in the intergenic region between *SEMA4A* and *SLC25A44* genes (chr1:156,147,543-156,163,730) (**Supplementary Figure S1**). Because this alignment failure affected the entire cohort, these two poorly covered regions were not included in subsequent steps of the analysis to reduce risk of technical bias. Excluding these low coverage regions, average read depth was > 290.

### 2. Single variant analyses prioritize variants in *SEMA4A* 5’-UTR and *PMF1* intronic regions

A combination of approaches was used to assess the association of single variants with risk of ICH at *1q22*. Firth regression analysis comparing cases (lobar and non-lobar, separately) and controls, was computed to test for association of the common variants with ICH. Given the minimal overlap with published GWAS analyses, our results replicate the non-lobar signal previously detected in proximity of *PMF1*^5^ (**Supplementary Figure S2A, B**), corroborating the role of this locus in risk for non-lobar ICH. When focusing on the magnitude of the effect (i.e., absolute value of the log of the odds ratio) rather than p-values alone, we identified an additional pool of variants within the *SEMA4A* 5’-UTR region. This signal was not previously reported because it did not reach genome-wide significance thresholds, but in review of the prior GWAS dataset these variants displayed a similarly elevated odds ratio (OR)^5^ (**Supplementary Figure S2A**,**B**). An additional small region with elevated ORs was detected in *SEMA4A* 3’-UTR region. Because variants located between base pairs 156,148,200 and 156,153,800 were entirely overlapping with the low coverage intergenic region previously mentioned, we did not consider them in subsequent analyses due to concerns for genotyping accuracy.

To improve our power, we performed a second analysis comparing non-lobar ICH cases against lobar ICH cases and controls pooled together, based on the prior observation that variants at *1q22* increase the risk of non-lobar ICH alone as well as the confirmation of no significant associations between lobar ICH and controls (**Supplementary Figure S2E**); that is, for the latter, the allele frequencies in lobar ICH cases were comparable to the controls. Because this yielded similar effect sizes with smaller standard errors, hence greater statistical power, compared to the non-lobar vs controls-only analysis (**Supplementary Figure S2B, C)**, we retained this analytic approach in all following analyses. Regressions performed adjusting, or not, for principal components produced similar results, excluding the presence of population stratification **(Supplementary Figure S2C**,**D)**. We note that examining the eQTLGen Consortium data for this region, the associated variants in this newly detected *SEMA4A* 5’-UTR were predicted to be *PMF1* eQTLs in blood (**Fig 2A**).

**Figure 2.**
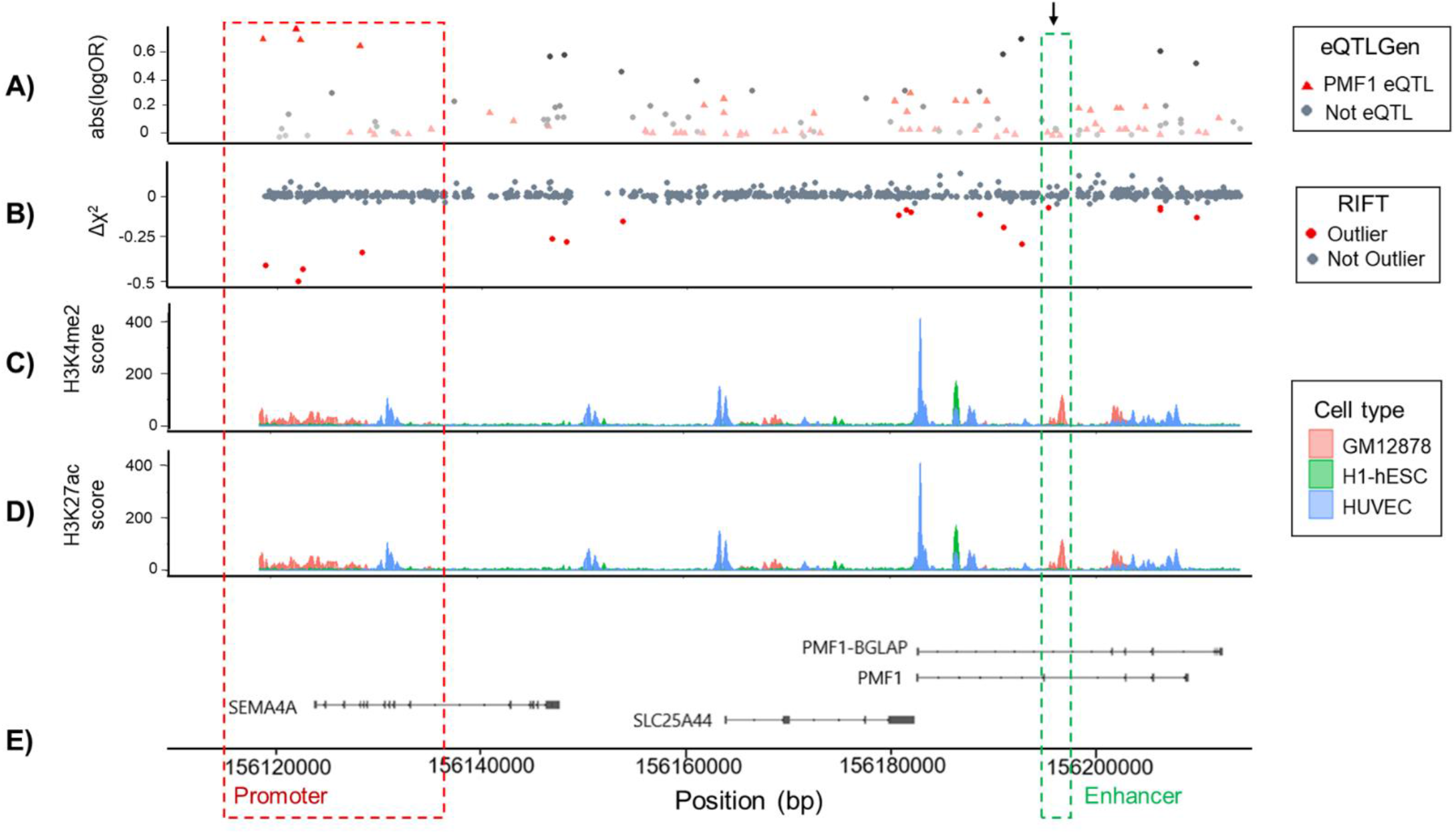
Single variant analysis and ENCODE/Ensembl annotation at *1q22*. A) Firth analysis results prioritize variants falling in proximity of *SEMA4A* 5’-UTR and *PMF1* intronic region. Absolute values of natural logarithm of Odds ratio and genomic position are plotted for each variant included in Firth regression analysis. Red triangles identify variants reported acting as *PMF1* eQTL in blood after eQTLGen data query, while grey dots represent variants not acting as *PMF1* eQTLs. An arrow indicates the location of the top hit previously prioritized by the GWAS analysis of non-lobar ICH performed by Woo *et al*.^5^ B) RIFT analysis identifies potentially causal variants falling in *SEMA4A* 5’-UTR and *PMF1* intronic regions. Delta-Chi square values resulting from RIFT analysis are plotted for each variant against together with its genomic position. Outliers are colored in red. C) Analysis of ENCODE H3K4me2 histone marks highlight that *1q22* is a transcriptionally active region. H3K4me2 score levels measured in GM12878, H1-hESC, HepG2 and NHLF cell types. Higher peaks indicate the presence of enrichment in transcription factor binding sites. D) ENCODE H3k27ac marks analysis points out the presence of active promoter and enhancer regions, as shown by high H3k27ac score signals. E) Red and green dashed rectangles define the regions identified as active promoter and enhancer following Ensembl database query.

We pursued our single rare variant analysis using RIFT,^13^ that prioritized variants in the two regions previously identified by Firth regression, the *SEMA4A* 5’-UTR and *PMF1* intronic region (**Fig 2B**).

We also used the Wald Z-scores from Firth regression to perform fine-mapping analysis combining functionally-informed and statistical fine-mapping approaches. Both methods further corroborated the results from Firth and RIFT analyses, prioritizing variants in *SEMA4A* 5’-UTR and *PMF1* intronic regions (**Supplementary Fig S3**).

### 3. Functional annotation identifies regions overlapping with active promoter and enhancer elements

We used ENCODE and Ensembl databases to investigate whether the two regions prioritized by the single variant analyses could have a functional role at *1q22*. H3K4me2 signal analysis showed enrichment in transcription factor binding sites in the proximity of both *SEMA4A* 5’-UTR and *PMF1* intronic regions (**Fig 2C**), suggesting the presence of active regulatory elements. This was further supported by H3K27ac marks, highlighting the presence of active promoter and enhancer regions overlapping with the prioritized *SEMA4A* 5’-UTR and *PMF1* intronic regions (**Fig 2D**). Ensembl database queries corroborated these results, classifying active *SEMA4A* 5’-UTR promoter (chr1:156,115,002-156,136,199) and *PMF1* enhancer (chr1:156,194,401-156,194,600) regions at these same locations (**Fig 2D**), in a variety of different cell types, including vascular cells and neurons.

Taken together, common and rare single variant analyses combined with functional annotation suggest a role for *SEMA4A* 5’-UTR promoter and *PMF1* enhancer regions in non-lobar ICH susceptibility through a mechanism of transcriptional regulation of genes at *1q22*.

### 4. *1q22* is spatially organized as a single transcriptionally active domain

We used publicly available Hi-C data via Juicebox to evaluate 3D chromatin organization at *1q22*. Juicebox representations demonstrated a major contact domain spanning a larger region surrounding *1q22* (chr1:156,045,001-156,305,000). Within this larger region, two smaller contact domains were detected, one of which comprised a TAD containing the *1q22* locus (chr1:156,115,001-156,200,000) (**Fig 3A**). Juicebox also localized two chromatin peaks at chr1:156,125,001-156,130,000 and chr1:156,190,001-156,195,000, suggesting the formation of a chromatin loop containing 1q22 (**Fig 3B**).

**Figure 3.**
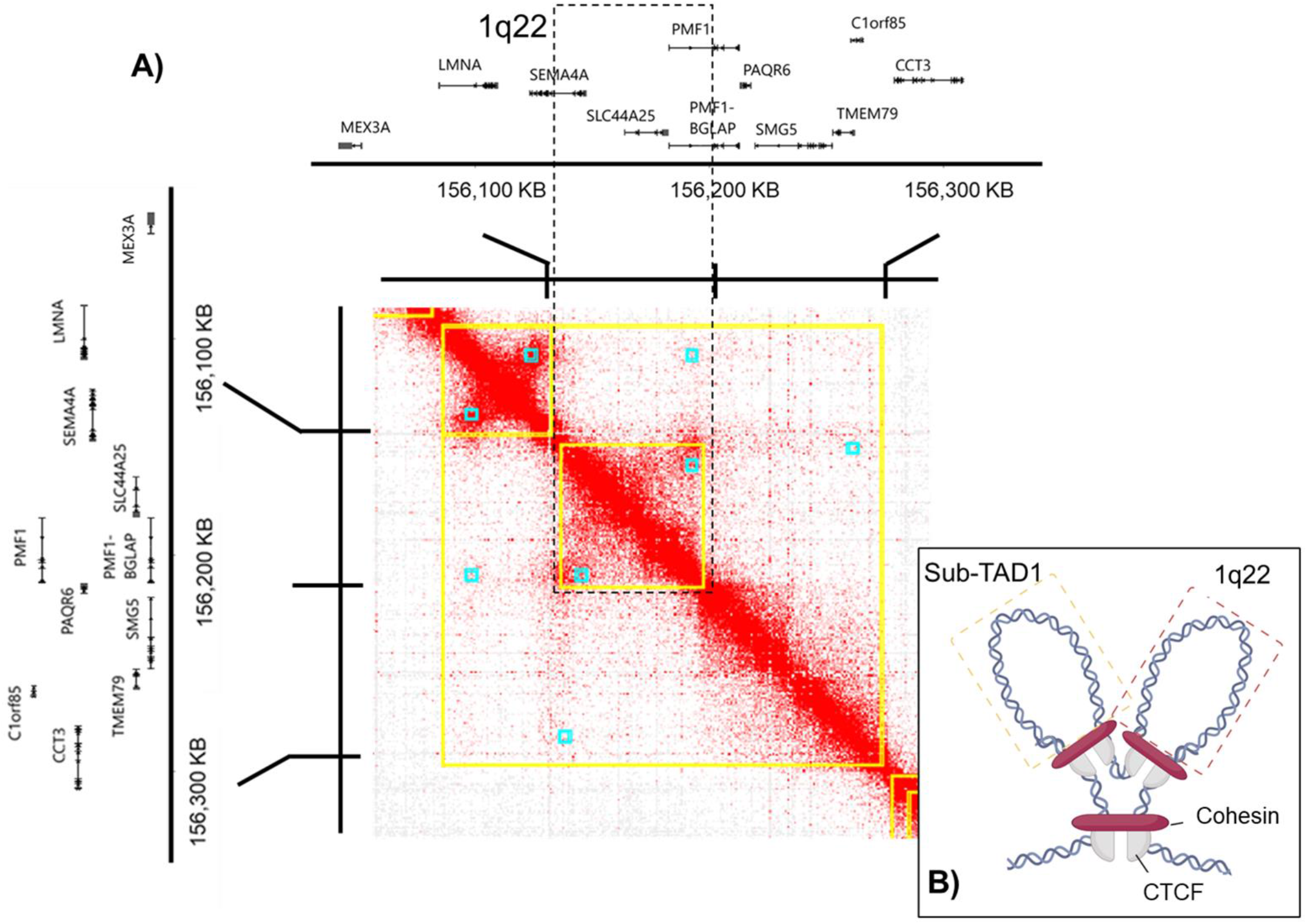
Hi-C data allow to explore *1q22* 3D chromatin conformation, highlighting that *1q22* is organized as a single TAD. A) Juicebox analysis highlights the presence of contact domains (yellow squares) and chromatin peaks (blue squares) within *1q22* and led us to hypothesize that the region surrounding the locus is organized as a major transcriptionally active domain (TAD) harboring two sub-TADs, one adjacent to *1q22* (sub-TAD 1) and one encompassing *1q22* (B).

Seeking additional evidence on the presence of physical interactions within *1q22*, we examined ENCODE ChIP-Seq and ChIA-PET data generated in K562 cells. ChIP-Seq data showed two CTCF peaks in proximity to the *SEMA4A* 5’-UTR and *PMF1* intronic regions (**Fig 4A**), indicating enrichment of CTCF binding sites to these two regions which is indicative of loop formation. Next, we examined ChIA-PET data for CTCF motifs and POLR2A binding sites to attempt to identify the presence of long-range looping interactions within *1q22*. These data highlighted the presence of CTCF (chr1:156,115,504-156,116,593 and chr1:156,194,906-156,195,948) and POLR2A binding regions (chr1:156,128,270-156,132,924 and chr1:156,182,023-156,185,783), further supporting the presence of long-range interactions at *1q22*.

**Figure 4.**
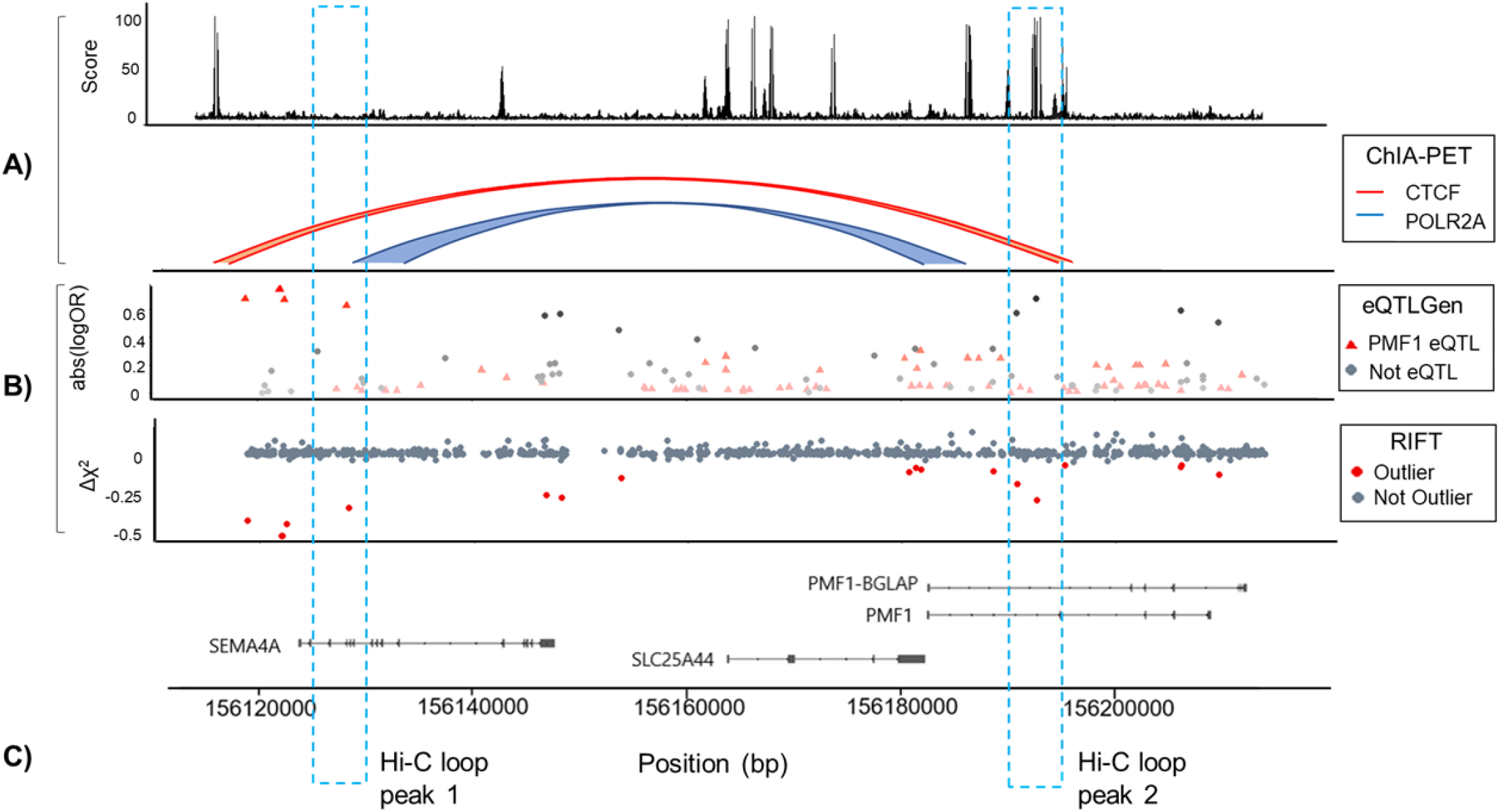
ENCODE ChIP-Seq and ChIA-Pet data analysis allow to identify the presence of long-range interactions within *1q22*. **A**) ChIP-Seq ad ChIA-Pet results highlight the presence of CTCF enrichment and long-range interactions within *1q22*. B) Variants previously prioritized by Firth and RIFT analysis fall within, or in close proximity, to the regions involved in long-range interactions. C) Dashed blue rectangles delimit the two regions interacting to form chromatin loops, as predicted by Juicebox.

Taken together, the analyses of *1q22* chromatin organization show that the genes belonging to this locus are within the same TAD, indicating that they are likely co-expressed. In addition, the combination of Hi-C, ChIP-Seq and ChIA-PET data analyses detected the presence of long-range interactions between the *SEMA4A* 5’-UTR promoter and *PMF1* enhancer regions previously prioritized (**Fig 4A**). Interestingly, all variants previously prioritized fell within or in close proximity of the two regions involved in long-range interactions and chromatin loop formation (**Fig 4B-C**). Thus, we hypothesized that variants modifying these interactions, as well as altering chromatin loop formation, could have an impact on *1q22* genes’ expression, potentially causing the higher non-lobar ICH risk observed at this locus.

### 5. *PMF1* over-expression is causally associated with higher non-lobar ICH risk at *1q22*

Building upon the evidence from chromatin conformation analyses that there is a TAD across the sequenced gene region at *1q22*, we assessed the causal role of the *1q22* genes on non-lobar ICH susceptibility, using variants associated with expression of those genes in multivariable Mendelian randomization analysis. Blood expression data released by the eQTLGen Consortium were screened to select genetic instruments associated with increased expression for each gene. Estimates from the GWAS analysis by Woo *et al*^5^ provided the effects of the selected genetic instruments on the outcome. MVMR was performed including variants influencing *SEMA4A, SLC25A44* and *PMF1* genes expression (*cis*-eQTLs data were not available for *PMF1-BGLAP* gene). Exposure and outcome data harmonization yielded a total of 13 variants that were used as instruments in subsequent MVMR analyses (**Supplementary Table 1**). Only *PMF1* overexpression was significantly associated with non-lobar ICH risk (**Fig 5**), suggesting a potential causal role within the base assumptions of the MVMR models used.

**Figure 5.**
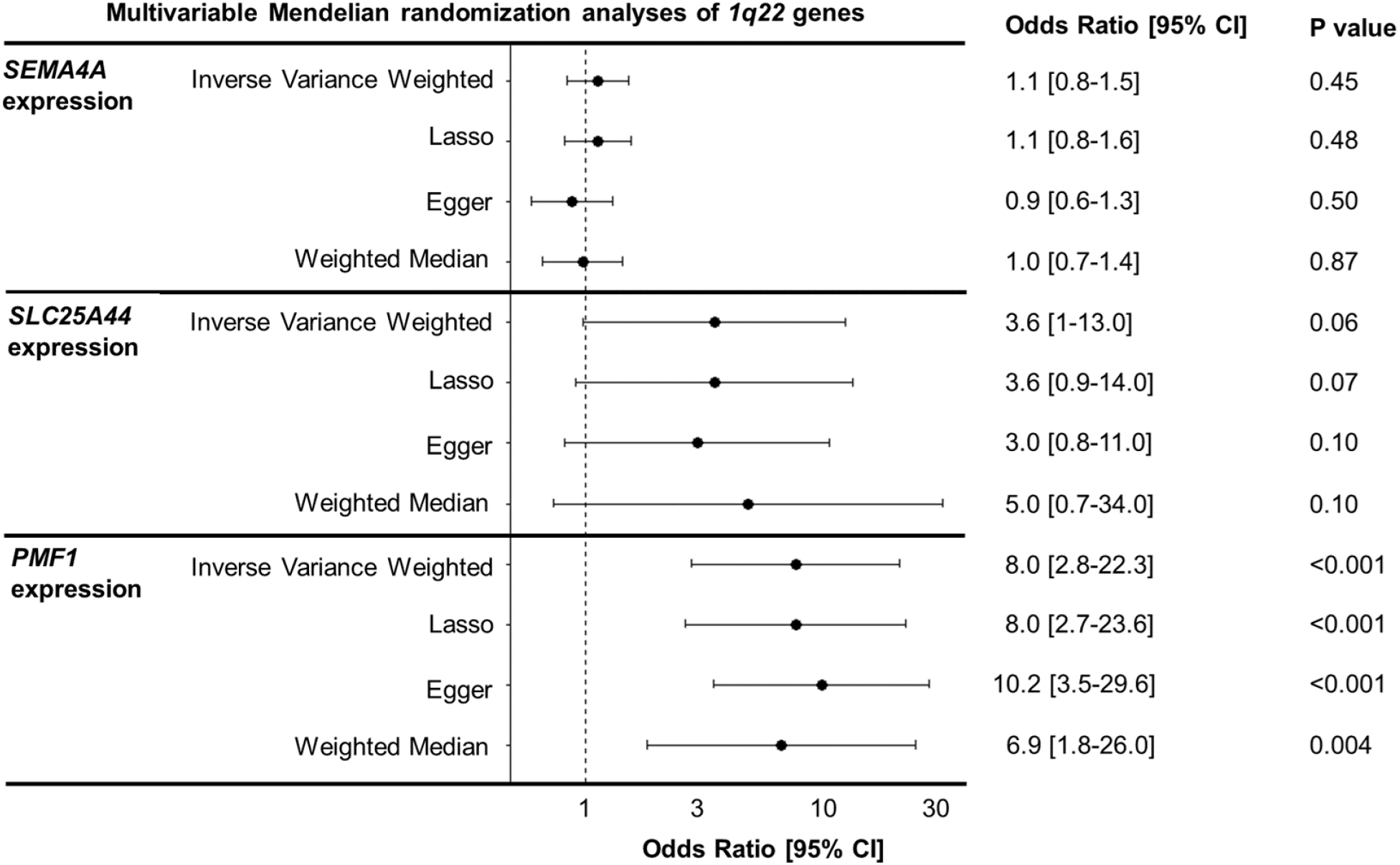
Multivariable Mendelian randomization analyses highlight that *PMF1* overexpression is causally associated to higher non-lobar ICH risk. Odd ratios, 95% confidence intervals, and p-values resulting from multivariable Mendelian randomization analyses are reported for each gene tested.

## Discussion

We sought to identify potential causal genetic mechanisms within the non-lobar ICH susceptibility *1q22* region identified in prior GWAS studies of ICH and related CSVD traits.^5–8^ After performing deep resequencing of the region, we identified common and rare variants in the *SEMA4A* promoter and *PMF1* enhancer regions associated with non-lobar ICH. Despite their physical distance, these associated variants in the *SEMA4A* promoter region were predicted to act as *PMF1* eQTLs, altering *PMF1* expression levels. Exploring epigenetic datasets to investigate the locus 3D organization, we identified evidence that *1q22* is spatially organized within a single chromatin loop and that the encoded genes belong to the same TAD, reflecting potentially shared expression regulation processes. The presence of long-range interactions involving the *SEMA4A* promoter and *PMF1* enhancer regions prioritized by the single variant analyses further support a shared gene expression regulatory process across these two regions and provide insight into the mechanism of how variants in the *SEMA4A* promoter region may act as *PMF1* eQTLs. The complementary MVMR analyses controlling for coexpression across *1q22* are consistent with a potential causal role for *PMF1* overexpression in mediating non-lobar ICH risk.

The identification of non-coding variants conferring disease susceptibility is common in GWAS studies.^34^ Such variants can express their pathogenic effect by altering expression regulation processes, as they modify promoter and enhancer activity.^35^ The increasing availability of public epigenetic datasets has enhanced the toolkit available for investigating non-coding variant effects, extending beyond eQTL libraries and into resources to explore 3D chromatin architecture as a means to develop testable hypotheses for biological investigation. The presence of variants capable of altering long-range chromosomal interactions, leading to transcriptional changes, has already been reported as a pathological mechanism in neurologic disorders such as schizophrenia, Alzheimer’s disease, and major depressive disorder.^34,36^ An elegant demonstration of this pathological mechanism was provided by a recent study that dissected a GWAS locus for frontotemporal lobar degeneration, showing that the risk haplotype is associated with increased CTCF recruitment and chromatin loop formation, leading to overexpression of *TMEM106B* and ultimately cytotoxicity.^36^ This demonstrated role of alteration in chromatin architecture as a pathogenetic mechanism in the etiology of sporadic neurological diseases supports our observations in ICH, with variants altering long-range interactions between *1q22* promoter and enhancer regions potentially leading to dysregulation of *PMF1*.

Encoding for polyamine modulating factor-1, PMF1 plays an active role in the catabolic pathway of polyamine metabolism. Polyamines (putrescine, spermidine and spermine) are essential for normal cellular growth and development, and activation of their catabolic pathway has been linked with tissue damage associated with pathological conditions, including stroke.^37^ PMF1 is directly involved in inducing the transcription of spermidine/spermine-N(1)-acetyltransferase (SSAT), one of the pathway’s rate-limiting enzymes,^38^ essential to prevent polyamine accumulation.^38^ This preventive pathway occurs at the expense of reactive oxygen species (ROS) production, as well as of other potential toxic metabolic byproducts, such as acrolein,^39^ a highly reactive compound reported to be more toxic than common ROS.^40^ Cytotoxic and neurotoxic effects of acrolein have been extensively reported, and high acrolein levels have been proposed to mediate pathological mechanisms underlying brain infarction in mouse models,^41–43^ as well as blood vessel rupture in cellular models.^44–47^ Elevated levels of acrolein and SSAT have also been detected in the plasma of stroke patients and have been proposed as candidate stroke biomarkers.^48–50^ Overall, this prior evidence contextualizes our observations at *1q22*, and provides some clues as to how dysregulation of a gene such as *PMF1* might lead to a complex disease like ICH. A testable hypothesis supported by our observations and these previous studies is that *PMF1* overexpression caused by chromatin loop alteration could enhance SSAT transcription. Higher SSAT levels could, in turn, increase acrolein production and consequent accumulation. High acrolein levels could then induce oxidative stress and exert a cytotoxic effect by increasing small vessels’ susceptibility to injury, ultimately resulting in non-lobar ICH. While additional research is clearly needed to further probe this mechanism and understand why it appears to be less relevant to the lobar ICH phenotype, we posit that our results provide a lens to help focus downstream *in vitro* and *in vivo* work focusing on the role of *PMF1* in non-lobar ICH.

The combination of multiple complementary methods empowered by a variety of publicly available datasets greatly facilitated our exploration of the genetic architecture of *1q22* in non-lobar ICH, with implications that extend to other CSVD phenotypes that share this susceptibility locus. Collectively, our results support a role for *PMF1* as the mediator of non-lobar ICH observed at this locus, potentially acting through its known role in polyamine regulation which would represent a novel therapeutic target. However, our work has several limitations. First, limited statistical power to discover or rediscover variants reaching genome-wide significance led to nominal statistical associations at the single variant level, motivating much of our study design. Additional follow-up studies, including larger and multi-ancestral sample sizes and more narrowly refined regions motivated by this research will be fundamental to confirm our findings at *1q22* and identify variants or haplotypes potentially implicated in enhancing or repressing loop formation. While publicly available ChIP-Seq and ChIA-PET datasets were crucial in corroborating our observations, dedicated epigenetic work focused on the *1q22* locus is needed to replicate and extend our findings. While MVMR can support causal directional associations in the appropriate context, its results must be tested in future transcript and protein-level experiments. Engineered cell lines harboring specific variants in the *SEMA4A* 5’-UTR promoter and *PMF1* enhancer regions are likely to be useful in furthering several of the testable hypotheses arising from this work, building towards novel therapeutic targets for ICH and related common diseases of the cerebral small vessels for which no specific treatment currently exists.

## Data Availability

Additional data supporting these findings are available by the authors, upon reasonable request.

https://www.ncbi.nlm.nih.gov/gap/

https://cd.hugeamp.org/downloads.html

https://www.eqtlgen.org

## Acknowledgments

This work is supported by R01NS103924, R01NS059727, R01NS100178, R01NS105150, U01NS069763 and U19NS115388. Targeted sequencing services were provided by the Northwest Genomics Center at the University of Washington, Department of Genome Sciences, under U.S. Federal Government contract number HHSN268201100037C from the National Heart, Lung, and Blood Institute (RS&G 224). Computations at Wake Forest were performed using the Wake Forest University (WFU) High Performance Computing Facility, a centrally managed computational resource available to WFU researchers including faculty, staff, students, and collaborators (URL https://doi.org/10.57682/g13z-2362).

## Author Contributions

L.P., M.E.C., C.D.L. and C.D.A. contributed to the conception, design and analysis of the study. All authors contributed to data acquisition. All authors contributed to manuscript drafting, revision and figures preparation.

