## Supplemental material for "Deep resequencing of the 1q22 locus in non-lobar intracerebral hemorrhage"

**Supplementary Material**

**Supplementary Figure S1.** *1q22* coverage analysis results. Average reads depth was calculated for each sample at each sequenced position. All the samples were characterized by low coverage, detected in a 1.6 kb region falling in an intronic region of *SEMA4A* gene (chr1:156,139,280 - 156,140,732) and in an intergenic region between *SEMA4A* and *SLC25A44* genes(chr1:156,147,543-156,163,730). Since affecting the entire cohort, it was considered as a technical bias.

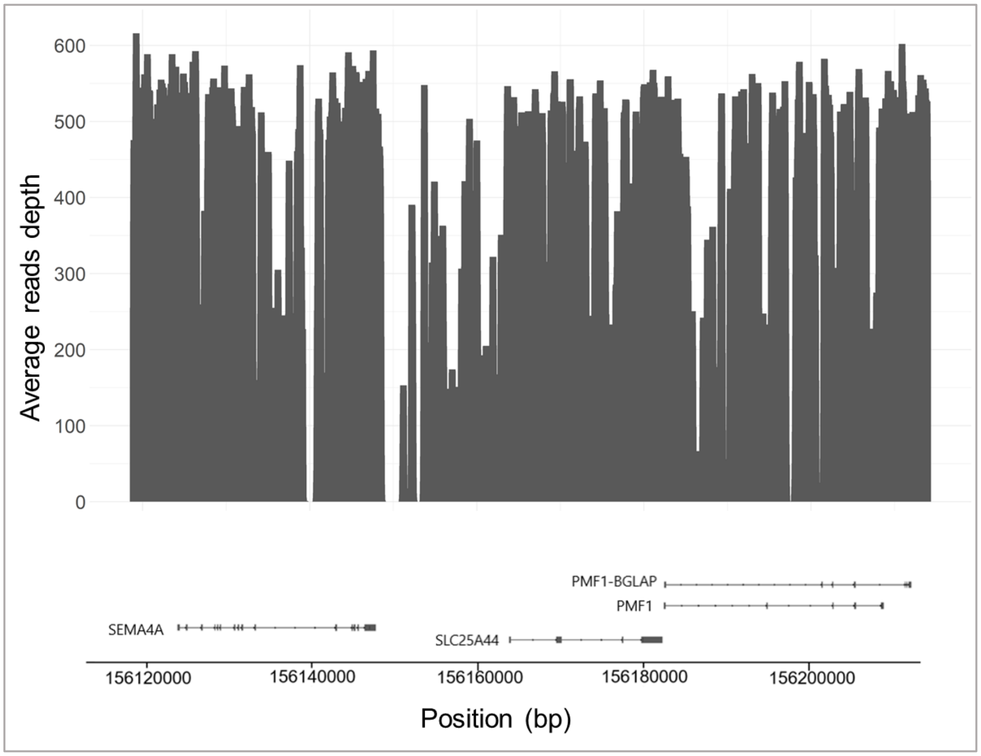

**Supplementary Figure S2**. P-values (left panel) and absolute abs(logOR) (right panel) of non-lobar ICH vs controls in meta-analysis by Woo et al (A) were compared with those resulting from Firth regression comparing non-lobar vs controls (B), non-lobar vs lobar and controls (C), non-lobar vs lobar and controls, adjusted for population stratification (D), lobar vs controls (E).

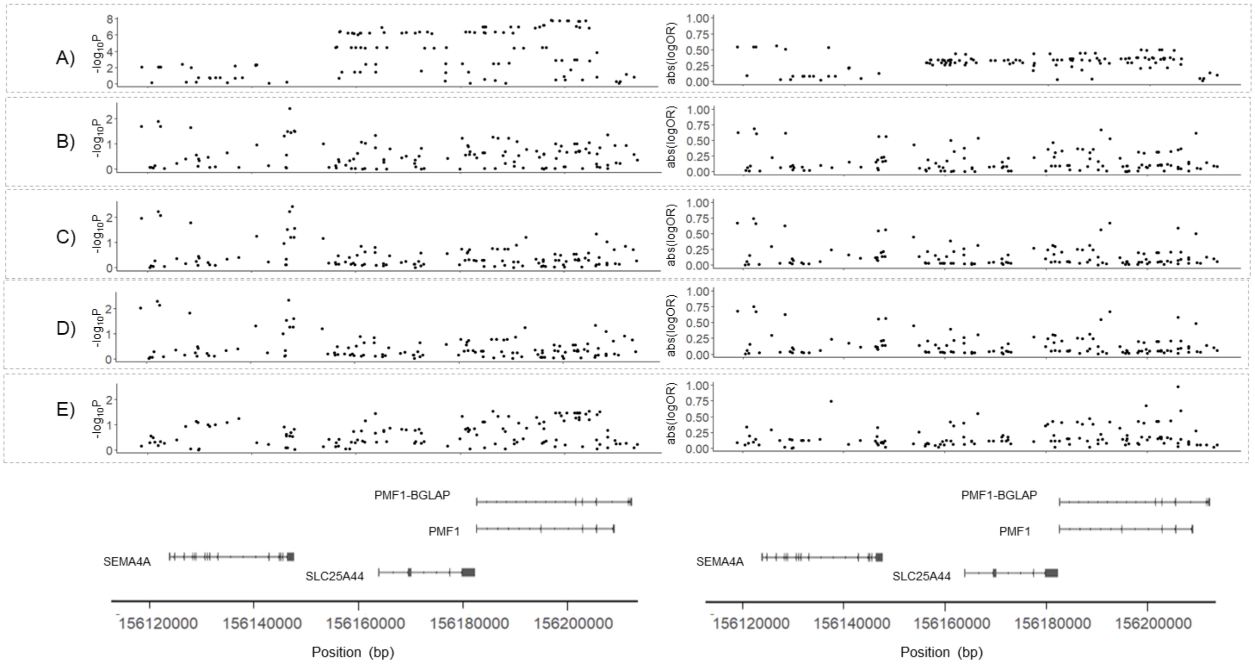


**Supplementary Figure S3**. Fine-mapping results. Posterior Inclusion Probabilities (PIPs) are reported for PAINTOR (A), SuSIE (B), FINEMAP (C) and Caviar (D) analyses.

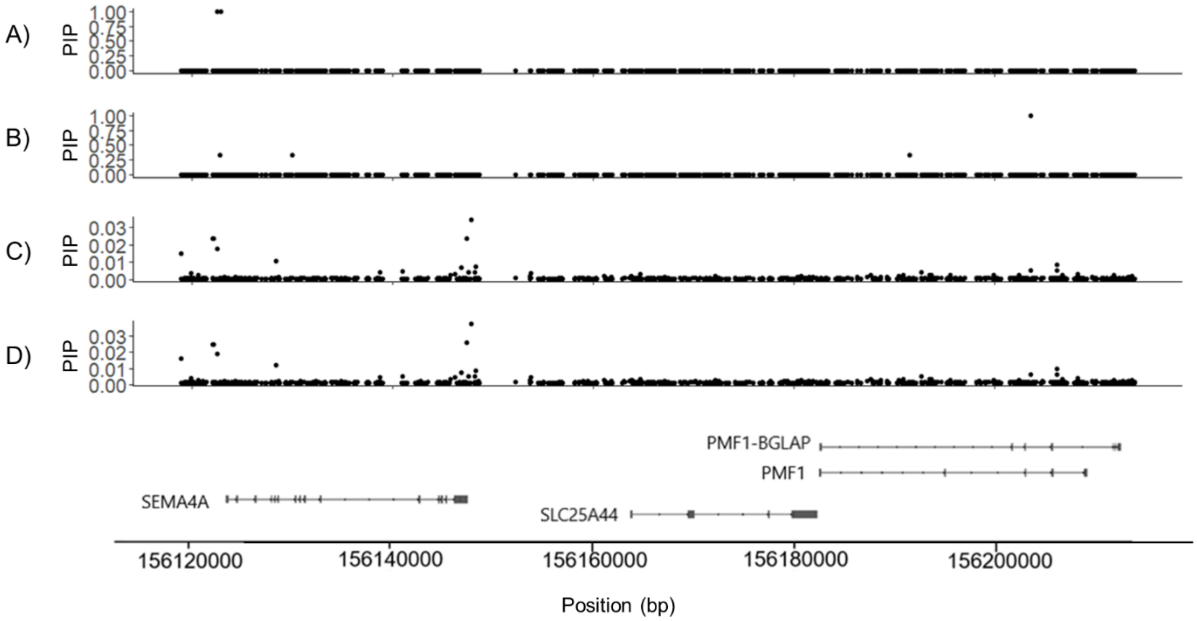

**Supplementary Table 1.** Variants used as genetic instruments in MVMR analyses are reported with correspondent betas (relative to effect alleles) and standard errors (SE).

| **SNP** | **Effect allele** | **Other allele** | **Beta nonlobarICH** | **SE nonlobarICH** | **Beta *SEMA4A*** | **SE *SEMA4A*** | **Beta *SLC25A44*** | **SE *SLC25A44*** | **Beta *PMF1*** | **SE *PMF1*** |
|---|---|---|---|---|---|---|---|---|---|---|
| rs112223391 | A | G | -0.29 | 0.07 | 0.35 | 0.01 | -0.04 | 0.01 | -0.12 | 0.01 |
| rs1543294 | T | C | 0.04 | 0.07 | -0.01 | 0.01 | -0.06 | 0.01 | 0.08 | 0.01 |
| rs2241106 | G | C | 0.06 | 0.07 | 0.30 | 0.01 | 0.01 | 0.01 | 0.02 | 0.01 |
| rs56410778 | C | T | -0.04 | 0.23 | 0.38 | 0.03 | -0.03 | 0.03 | -0.15 | 0.04 |
| rs72710255 | G | A | 0.08 | 0.09 | 0.10 | 0.01 | 0.01 | 0.01 | 0.02 | 0.01 |
| rs4661035 | G | A | -0.04 | 0.23 | 0.19 | 0.03 | 0.00 | 0.03 | -0.04 | 0.03 |
| rs16837430 | A | T | -0.01 | 0.09 | -0.07 | 0.01 | -0.02 | 0.01 | 0.00 | 0.01 |
| rs73006728 | A | G | -0.54 | 0.21 | -0.15 | 0.02 | -0.07 | 0.02 | -0.07 | 0.03 |
| rs2985714 | A | G | 0.25 | 0.10 | 0.08 | 0.01 | 0.00 | 0.01 | 0.02 | 0.02 |
| rs28662267 | C | T | -0.08 | 0.07 | -0.05 | 0.01 | 0.00 | 0.01 | 0.00 | 0.01 |
| rs2075163 | G | T | 0.02 | 0.18 | 0.12 | 0.02 | -0.08 | 0.02 | -0.04 | 0.02 |
| rs72704176 | T | C | -0.11 | 0.24 | -0.18 | 0.03 | -0.06 | 0.03 | -0.03 | 0.04 |
| rs1800247 | C | T | -0.13 | 0.07 | 0.01 | 0.01 | -0.07 | 0.01 | -0.04 | 0.01 |
